# Epigenetic clock acceleration is linked to earlier onset and phenoconversion age in REM sleep behavior disorder

**DOI:** 10.1101/2023.03.24.23287635

**Authors:** Konstantin Senkevich, Amélie Pelletier, Christine Sato, Allison Keil, Ziv Gan-Or, Anthony E. Lang, Ronald Postuma, Ekaterina Rogaeva

**Affiliations:** The Neuro (Montreal Neurological Institute-Hospital), McGill University, Montreal, Quebec, Canada; Department of Neurology and neurosurgery, McGill University, Montréal, QC, Canada, Canada; Tanz Centre for Research in Neurodegenerative Diseases, University of Toronto, 60 Leonard Ave., Toronto, ON, M5T 0S8, Canada; Center for Advanced Studies in Sleep Medicine, Montreal Sacre Coeur Hospital, Montreal, Quebec, Canada; Department of Human Genetics, McGill University, Montréal, QC, Canada; Edmond J. Safra Program in Parkinson’s Disease, Toronto Western Hospital, Toronto, Ontario, Canada; Morton and Gloria Shulman Movement Disorders Clinic, Toronto Western Hospital, Toronto, Ontario, Canada; Division of Neurology, Department of Medicine, University of Toronto, Toronto, Canada; The Research Institute of the McGill University Health Centre, Montreal, Quebec, Canada

## Abstract

Rapid-eye movement sleep behavior disorder (RBD) is the strongest prodromal marker for α-synucleinopathies. The Horvath DNA-methylation-age (DNAm-age) is an epigenetic clock that could reflect biological aging. We evaluated whether RBD age-at-onset/phenoconversion are associated with DNAm-age-acceleration in 162 RBD patients. We found an association of DNAm-age-acceleration with RBD age-at-onset at baseline (P=2.59e-08) and at follow-up (N=45; P=9.73e-06). RBD patients with faster aging had 4.6 years earlier onset than patients with slow/normal aging. Similarly, earlier age-at-phenoconversion was associated with DNAm-age-acceleration (N=53; P=1.26e-04). We demonstrated that epigenetic clock acceleration is linked with an earlier RBD age-at-onset and, hence with earlier phenoconversion.

## Introduction

Rapid-eye movement (REM) sleep behavior disorder (RBD) is a condition in which individuals act out vivid and often violent dreams during REM sleep.^1^ RBD is the strongest known prodromal marker for α□synucleinopathies, including Parkinson’s disease (PD), Lewy Body Dementia (LBD) and Multiple system atrophy (MSA).^2, 3^ RBD is linked to a more severe subtype of α□synucleinopathy, with a higher chance of developing dementia or other non-motor symptoms after phenoconversion.^4^ The annual conversion rate of RBD to PD-related syndromes was reported to be 6.3%, reaching 74% after 12-year follow-up.^5^

There is high phenotypic heterogeneity in RBD, including a wide range in age-at-onset or age-at-phenoconversion to PD, LBD or MSA (ranging from 2 to 12 years from diagnosis).^5^ However, knowledge about disease modifiers is limited. The variable phenotypes and age-at-phenoconversion could be linked to epigenetic mechanisms affecting downstream protein levels and cellular function.^6^ Epigenetic mechanisms are central to neurodevelopment, synaptic transmission, and plasticity; and have been implicated in numerous brain illnesses, including PD.^7, 8^ One of the key epigenetic modifications is DNA methylation (DNAm) of cytosines at CpG-sites, which can be affected by environmental and genetic factors. Importantly, DNAm is closely linked to aging, the strongest risk factor for all neurodegenerative disorders.^9^

The aging process is variable between individuals and not well-reflected by chronological (calendar) age. Hence, efforts have been made to develop biomarkers of biological aging, the pace of which varies amongst individuals and reflects the functional capability of a person. The number of age predictors based on DNAm profile is rising due to their potential in predicting healthspan,^9^ including the multi-tissue Horvath clock consisting of ∼350 CpGs (hyper[or hypo-methylated with age). The cumulative assessment of these CpGs allows for the estimation of DNAm-age, which is an accurate age predictor across different tissues, including blood and brain.^10^

The difference between DNAm-age and chronological age (DNAm-age-acceleration) is associated with age-at-onset of amyotrophic lateral sclerosis (ALS) and PD.^11, 12^ We also reported a large family with the p.A53E mutation in α□synuclein in which an earlier PD onset was accompanied by increased DNAm-age-acceleration.^13^ However, it is not known whether RBD age-at-onset and age of phenoconversion of RBD to an overt clinical α□synucleinopathy are associated with DNAm-age-acceleration, which were evaluated in the current study.

## Methods

### Population

Ethics approval for the study was obtained from the local research ethics board and all study participants signed informed consent. The diagnosis of polysomnography-confirmed idiopathic RBD was made based on the standard International Classification of Sleep Disorders criteria.^14^ Onset age was defined by patient self-report. Our cohort included 162 RBD patients, a large part of which (N=154) has been comprehensively described elsewhere.^5^ A longitudinal assessment was available for 45 patients, who had donated blood at two time-points (2-10 years apart). Patients were assessed annually (RP) for clinical signs of α□synucleinopathy and 53 patients in our cohort phenoconverted to PD, LBD or MSA (Table 1).

**Table 1.** Demographics of the RBD cohort

|  | Time point |  |  |  |
| --- | --- | --- | --- | --- |
|  | Baseline patients (N=162) | Follow-up patients (N=45) | Phenoconverters (N=53)* |  |
|  |  |  | PD (N=30) | DLB (N=22) |
| Males (n, %) | 125, 77% | 38, 76% | 24, 80% | 15, 68% |
| Age at RBD onset, (Mean ± SD) | 56.42 ± 10.95 | 55.67 ± 9.92 | 55.00 ± 7.82 | 60.95 ± 13.22 |
| Age at diagnosis, (Mean ± SD) | 64.34 ± 7.76 | 63.44 ± 7.29 | 62.46 ± 6.30 | 69.64 ± 7.15 |
| Age at sample collection, (Mean ± SD) | 66.2 ± 8.15 | 69.33 ± 8.37 | 65.04 ± 7.50 | 72.73 ± 6.61 |
| DNA-methylation age (Mean ± SD) | 72.48 ± 7.46 | 74.23 ± 7.46 | 72.48 ± 6.61 | 76.28 ± 5.77 |
| Age-at-phenoconversion (Mean ± SD) | - | - | 68.63 ± 6.82 | 75.82 ± 6.43 |

\* – 1 patient phenoconverted to MSA (Male), SD – standard deviation, RBD – REM sleep behavior disorder, DLB – Lewy Body Dementia, PD – Parkinson's disease, MSA – multiple system atrophy

### DNAm analyses

Genomic DNA was extracted from blood samples using a QIAGEN kit and bisulfite converted using the EZ DNA Methylation-Lightning ™ Kit (Zymo), as reported previously.^15^ DNAm levels for age-related CpGs were obtained from the Infinium MethylationEPIC chip processed at the Microarray Facility at the Centre for Applied Genomics (Toronto). The β value was used to estimate the DNAm level of each CpG-site using the ratio of intensities between methylated and unmethylated alleles. Raw DNAm data was analyzed using the minfi package in R^16^ and processed by quantile normalization. We then extracted DNAm data of 334 age-related CpG-sites constituting the Horvath DNAm-age adjusted for blood cell counts based on the advanced analysis mode of the DNAm-age calculator (https://dnamage.genetics.ucla.edu/).^10^

### Statistics

DNAm-age-acceleration was calculated as DNAm-age minus chronological age at sample collection. We applied multivariate linear regression to estimate the association between DNAm-age-acceleration and either RBD age-at-onset or age-at-phenoconversion. The analyses were adjusted for sex and interval (the difference between age at sample collection and RBD onset). Taking under consideration the reported error for estimating DNAm-age of ∼3 years^10^, we also classified RBD patients into two groups: slow/normal aging (DNAm-age-acceleration <3 years) and fast aging (DNAm-age-acceleration >3 years). The hazard ratio (HR) with 95% confidence interval (CI) was calculated. P-value reported after adjustment for sex and interval.

## Results

### DNAm-age-acceleration is associated with age-at-onset of RBD

Multivariate linear regression analysis revealed an association between DNAm-age-acceleration with earlier RBD age-at-onset at baseline (N=162; B=-0.68; SE=0.12; P=2.59e-08; Figure 1A) and at follow-up in the group with a longitudinal assessment (N=45; B=-1.07; SE=0.21; P=9.73e-06; Figure 1B; Supplementary Figure 1). Average DNAm-age is presented in Table 1. Most of the RBD patients had fast epigenetic aging, defined as DNAm-age-acceleration >3 years (N=116; 72%), and those with fast aging had a 4.6-year earlier onset than patients with slow/normal aging (55.15±11.05 years vs 59.71±10.06 years). We also assessed whether there were any changes in DNAm-age-acceleration between the two time-points in the longitudinal cohort and did not observe any difference (5.77±5.53 at baseline vs 4.90±5.17 at follow-up; P=0.44).

**Figure 1.**
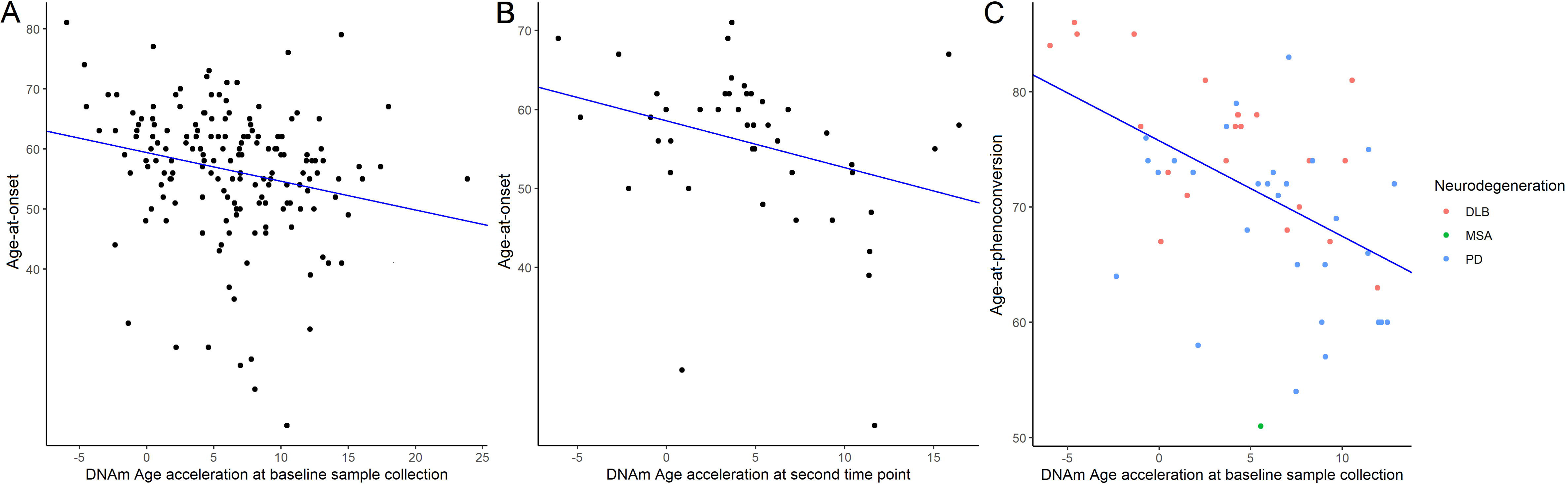
Scatter plot of DNAm-age-acceleration and age-at-onset or age-at-phenoconversion of RBD; A. at baseline (N=162; B=-0.68; SE=0.12; P=2.59e-08); B. at follow-up between 2-10 years (N=45; B=-1.07; SE=0.21; P=9.73e-06); and C. phenoconversion (N=53 B=-0.85; SE=0.21; P=1.26e-04). Linear regression analyses were adjusted for sex and interval.

### DNAm-age-acceleration is associated with age-at-phenoconversion of RBD

In our cohort, we had 53 phenoconverters to either PD (N=30), DLB (N=22) or MSA (N=1). During the prospective follow-up period, there was no association between DNAm-age-acceleration and risk of phenoconversion: 6.31±4.62 years in phenoconverters vs 6.23±5.27 in non-phenoconverters (P=0.92 on t-test; HR=1.12 (0.58–2,16), P=0.66). However, consistent with the results for age-at-onset, we found an association between DNAm-age-acceleration and age-at-phenoconversion (B=-0.85; SE=0.21; P=1.26e-04; Figure 1C), such that patients classified as faster aging had a 5.2-year earlier phenoconversion age (69.89±7.72 vs 75.06±7.96).

## Discussion

We found that DNAm-age-acceleration is associated with an earlier age of RBD onset and hence age of phenoconversion to clinically overt α□synucleinopathies, which is consistent with previous studies of neurodegenerative diseases (e.g., ALS and PD^11, 12^). Although we found no evidence that DNAm-age-acceleration is linked to risk of phenoconversion, this result could be affected by several factors. First, the prospective follow-up is relatively short compared to the total lifespan of an individual, so lack of power may be a key cause. Second, patients with younger RBD onset could have a different phenotype driving the earlier phenoconversion. Alternatively, once RBD has started, other factors (epigenetic or genetic) could affect phenoconversion independently from the epigenetic clock.

Environmental or genetic factors can affect DNAm and disrupt the epigenetic clock but the impact of DNAm-age-acceleration on neurodegenerative disorders is not well understood. Previous RBD studies have not investigated the relationship between DNAm and aging. However, reduced DNAm levels at specific genomic regions has been linked to RBD symptoms in PD patients with dementia, but not in DLB patients.^17^ Our study highlights the importance of further research into the underlying biological mechanisms of RBD, which could lead to improved diagnostic methods and treatments. DNAm-age could be an important predictor when designing a study or clinical trial to estimate rate of progression and define cut-off points. Moreover, recent randomized clinical trials demonstrated the response of the Horvath DNAm-age to diet and lifestyle,^18^ which encourage the use of the epigenetic clock to estimate the effectiveness of anti-aging interventions and serve as a potential therapeutic target.

Previously, a longitudinal analysis of PD progression revealed DNAm changes at several loci.^19^ Our RBD longitudinal analysis did not reveal a significant difference in DNAm-age-acceleration between the two time-points. This is consistent with a PD study showing that DNAm-age-acceleration was stable within a 3-year period for most carriers of *LRRK2* mutation, which supports that it could serve as a steady biomarker related to age-at-onset.^12^ Notably, a study including both identical and fraternal twins revealed high heritability of DNAm-age-acceleration in newborns (100%), and less heritability (but still noteworthy) in older subjects (39%) suggesting that non-genetic factors may gain greater relevance as individuals age.^10^

Our study has several limitations. First, age-at-onset of RBD is an estimate since it is self-reported and could be affected by other factors (e.g., whether the individual had a bed partner who noticed it earlier and how overt the dream enactment was). Second, we did not perform genetic stratification in our analysis, and future follow-up studies should assess large RBD cohort(s), including patients with genetic variants at *SNCA, GBA, TMEM175, INPP5F*, and *SCARB2*, which were associated with risk of RBD in a recent genome-wide association study.^20^ Third, our cohort is mainly of European origin and further studies in other populations will be required. Lastly, the aim of the study was to assess a link between DNAm-age-acceleration and RBD onset or phenoconversion. Therefore, we did not include controls, limiting our ability to detect specific DNAm regions affecting disease risk.

In conclusion, our findings suggest that epigenetic clock acceleration is a potential biomarker for earlier RBD onset. The use of a genomic blood-based biomarker of biological aging may also be valuable in assessing the effectiveness of anti-aging interventions in RBD, especially in clinical trials targeting modification of neurodegeneration or prevention of neurodegeneration onset. Further research, including replication of our results in other cohorts is needed to fully understand the link between DNAm-age and neurodegeneration, and to determine if DNAm-age-acceleration could be a target for future therapies.

## Data Availability

All results are reported in the tables or attached to the supplementary data.

## Acknowledgements

We would like to thank the participants for contributing to this study. This work was supported by the G. Harris Sheppard Fund for Research in Parkinson’s and other Neurodegenerative Diseases (ER), Canadian Consortium on Neurodegeneration in Aging (ER, ZGO), the Blidner Family Foundation (NPV), and the McLaughlin Accelerator Grants in Genomic Medicine (ER, ZGO, AEL, RP). ZGO is supported by the Fonds de recherche du Québec-Santé (FRQS) Chercheurs-boursiers award, in collaboration with Parkinson Quebec, and is a William Dawson Scholar. KS is supported by a post-doctoral fellowship from the Canada First Research Excellence Fund (CFREF), awarded to McGill University for the Healthy Brains for Healthy Lives initiative (HBHL), FRQS post-doctoral fellowship and McKerracher Award.

## Author Contributions

Concept and design: KS, AEL, RP, ER

Data acquisition: KS, AP, CS, AK, ZGO, AEL, RP, ER Statistical analysis: KS, AP, CS, AK, ZGO, AEL, RP, ER Drafting of the manuscript: KS, CS, AEL, RP, ER Critical revision of the manuscript: All

