## Supplementary Figure 1 for "Epigenetic clock acceleration is linked to earlier onset and phenoconversion age in REM sleep behavior disorder"

DNAm-age acceleration

| Years after baseline | R022 | R030 | R032 | R036 | R053 | R086 | R090 | R095 | R101 | R110 | R111 | R128 | R143 | R145 | R149 | R152 | R162 | R168 | R171 | R172 |
| --- | --- | --- | --- | --- | --- | --- | --- | --- | --- | --- | --- | --- | --- | --- | --- | --- | --- | --- | --- | --- |
| 0 | 1.24762 | 7.07595 | 0.349347 | 9.76653 | -1.21016 | 2.1851 | 4.16217 | 1.76585 | 4.28952 | 0.57553 | 12.2934 | 8.06683 | -2.21578 | 8.05256 | 5.34178 | 4.82708 | 5.9189 | 5.68635 | -0.7696 | 5.9799 |
| 4 |  |  |  |  |  |  |  | 4.83924 |  | 3.64236 |  | 11.6857 |  |  |  |  |  |  |  |  |
| 5 | 0.228157 |  |  |  |  |  |  |  |  |  |  |  | -6.07229 | 7.27873 |  |  |  | 4.03699 | 3.29534 | 3.66799 |
| 6 |  |  |  |  | -0.465233 |  |  |  |  |  |  |  |  |  | 3.51817 | 3.43563 | 5.40229 |  |  |  |
| 7 |  |  |  | 4.74945 |  |  |  |  |  |  |  |  |  |  |  |  |  |  |  |  |
| 9 |  |  |  |  |  |  | 9.31871 |  | 4.8941 |  |  |  |  |  |  |  |  |  |  |  |
| 10 |  | -0.882871 | 1.23571 |  |  | 0.873487 |  |  |  |  | 5.6934 |  |  |  |  |  |  |  |  |  |

Supplementary Figure 1. Change in DNAm-age-acceleration for the longitudinal samples (N=45) between the two time-points (2-10 years). Each different coloured line represents a different patient.
